# Logistic advantage of two-step screening strategy for SARS-CoV-2 at airport quarantine

**DOI:** 10.1101/2021.01.25.21250509

**Authors:** Isao Yokota, Peter Y Shane, Takanori Teshima

**Author notes:** Correspondence to: Dr Isao Yokota, Department of Biostatistics, Hokkaido University Faculty of Medicine, N15, W7, Kita-ku, Sapporo, 060-8638, Japan, and Prof Takanori Teshima, Department of Hematology, Hokkaido University Faculty of Medicine, N15, W7, Kita-ku, Sapporo, 060-8638, Japan.

## Abstract

**Background:** Airport quarantine is required to reduce the risk of entry of travelers infected with severe acute respiratory syndrome coronavirus 2 (SARS-CoV-2). However, it is challenging for both high accuracy and rapid turn-around time to coexist in testing; polymerase chain reaction (PCR) is time-consuming with high accuracy, while antigen testing is rapid with less accuracy.

**Methods:** 88,924 (93.2%) of 95,457 arrivals at three international airports in Japan were tested for SARS-CoV-2 using self-collected saliva by a screening strategy with initial chemiluminescent enzyme immunoassay (CLEIA) followed by confirmatory nucleic acid amplification tests (NAAT) only for intermediate range antigen concentrations.

**Results:** 254 (0.27%) persons were found to be SARS-CoV-2 antigen positive (≥ 4.0 pg/mL) by CLEIA. NAAT was required for confirmatory testing in 513 (0.54%) persons with intermediate antigen concentrations (0.67-4.0 pg/mL) whereby the virus was detected in 34 (6.6%) persons. This two-step strategy dramatically reduced the utilization of NAAT to approximately one out of every 200 test subjects.

Estimated performance of this strategy did not show significant increase in false negatives as compared to performing NAAT in all subjects. Further reduction in imported cases may be achieved by post-screening quarantine.

**Conclusions:** Point of care testing by quantitative CLEIA using self-collected saliva is less labor-intensive and yields results rapidly, thus suitable as an initial screening test. Reserving NAAT for CLEIA indeterminate cases may prevent compromising accuracy while significantly improving the logistics of administering mass-screening at large venues.

## Introduction

The coronavirus disease-19 (COVID-19) pandemic has forced many countries to introduce border closures to prevent the entry of infected travelers from regions where COVID-19 is rife^1-3^. However, this decision has brought on heavy social and economic repercussions. Now several countries have been increasingly accepting international flights, albeit with quarantine for any traveler suspected of infection by various tests^4-7^. Testing measures include PCR before departure, temperature and symptom check at airports, and PCR testing at arrival.

Due to the high volume of international air travel, performing the virus tests with high accuracy in rapid turnaround time has been challenging. Currently, the “gold standard” of severe acute respiratory syndrome coronavirus 2 (SARS-CoV-2) detection is nucleic acid amplification tests (NAAT), such as the quantitative reverse transcriptase polymerase chain reaction (PCR) using nasopharyngeal swab (NPS) samples^8, 9^. Recently, self-collected saliva has attracted attention as an alternative sample with significant logistic advantages over NPS^10^. Specifically, self-collection eliminates the requirement of specialized medical personnel and allows simultaneous parallel sample collection thus more suitable for mass screening^9, 11^. We and others have shown that the accuracy of self-collected saliva and NPS samples are equivalent in large scale direct comparative studies ^12-14^. However, although PCR is highly accurate and reliable, results may take 24-48 hours to return. Such delays may lead to further transmission of disease^15^, especially in the confines of airport transit. Recently, we and others have shown the utility of a novel quantitative antigen test using chemiluminescent enzyme immunoassay (CLEIA) and reverse transcriptase loop-mediated isothermal amplification (LAMP) to detect SARS-CoV-2 proteins and nucleic acids, respectively, within 30 minutes^14, 16^. Accordingly, a novel two-step strategy using these tests have been implemented for mass screening of SARS-CoV-2 at airport quarantines in Japan^17^. We herein evaluated utility of this strategy in 88,924 persons and estimated its performance in several clinical scenarios.

## Methods

### Design and Population

Testing for SARS-CoV-2 using either self-collected saliva or NPS samples obtained by medical officers was mandatory for all international arrivals between July 29 and September 30, 2020. Due to logistical advantages, vast majority of tests were performed using self-collected saliva. Samples were collected in a sterilized 15mL polystyrene sputum collection tube (Toyo Kizai, Warabi, Japan) and all specimens were analysed at the airport quarantine laboratories. This study was approved by the Institutional Ethics Board (Hokkaido University Hospital Division of Clinical Research Administration Number: 020-0116) and anonymously processed data were provided by the quarantine stations.

### Two-step strategy using CLEIA and NAAT

Initially, all specimens were tested by CLEIA with positive and negative thresholds of 4.0pg/mL and 0.67pg/mL, respectively, as previously reported^17^. Concentrations in between the thresholds were considered indeterminate, and only these specimens underwent confirmatory testing by NAAT.

### CLEIA

Lumipulse SARS-CoV-2 Ag kit (Fujirebio, Tokyo, Japan), a sandwich CLEIA using monoclonal antibodies that recognize SARS-CoV-2 N-Ag on LUMIPULSE G1200 automated machine (Fujirebio), was used as previously described^17^. The detection range is between 0.01 pg/mL and 5000 pg/mL.

### NAAT

Saliva was diluted 4-fold with phosphate buffered saline and centrifuged at 20,000 × g for 5 min to remove cells and debris. RNA was extracted from 200 µL of the supernatant using QIAsymphony DSP Virus/Pathogen kit and QIAamp Viral RNA Mini Kit (QIAGEN, Hilden, Germany). Then, nucleic acids of SARS-CoV-2 were detected by either PCR or LAMP. PCR tests were performed as previsouly described^14^. The cycle threshold (Ct)-values were obtained using N2 primers (NIID_2019-nCOV_N_F2, NIID_2019-nCOV_N_R2) and a probe (NIID_2019-nCOV_N_P2). LAMP was carried out to detect SARS-CoV-2 RNA using Loopamp® □ 2019-SARS-CoV-2 Detection Reagent Kit (Eiken Chemical, Tokyo, Japan) and the Loopamp Real-time Turbidimeter (Eiken Chemical) as previously described^14^.

### Statistical analysis

We compared the estimated effectiveness of three mass-screening strategies at border quarantine: the two-step strategy, NAAT only for all entrants (without CLEIA), and test-free entry, expressed as the rate of false negatives per 100,000 persons and the number of NAATs performed. The rate of false negatives by NAAT was estimated by *p* × *FN/Pos* where *p* is the assumed proportion of the test population with positive NAAT, and *FN/Pos* defined as the ratio of false negatives to all positives (i.e. the ratio of undetected infectees to persons diagnosed as infected). Four scenarios with *p* values of 0·1%, 0·2%, 0·5%, and 1·0% were used for analyses, with a factor of 0.76 (probability of CLEIA positivity given NAAT positivity) applied to the *p* values for the two-step strategy, consistent with our previous report^17^. *FN/Pos* was set at 0·4 in accordance to a recent report showing 136 and 52 positive results at airport screening and during post-entry compulsory quarantine, respectively^18^. Additional analyses were performed for *FN/Pos* set at 1.0 and 2.0. The impact of 14-day quarantine was calculated in all cases as a linear variable of adherence rates with all false negatives becoming apparent with adherence rate of 100%. By expressing *a* as the probability of CLEIA-positive given NAAT-positive, the rate of false negatives by CLEIA may be calculated as *p* × [*FN/Pos* + (1 - *a*)]. All statistical analyses were conducted by R 4·0·2 (R Foundation for Statistical Computing, Vienna, Austria).

## Results

88,924 persons screened by the two-step strategy using self-collected saliva accounted for 88.0% of all arrivals to Japan during the period. Initial CLEIA was found to be positive in 254 (0.29%) persons (Fig. 1). The 513 samples (0.58%) with antigen concentrations in the indeterminate range (between 0·67 pg/mL and 4·00 pg/mL) proceeded to NAAT with 34 (6.6%) positive results. 254 (99.2%) of the 288 positive results were diagnosed by the initial CLEIA, with only 34 (11·8%) diagnosed by NAAT. On the other hand, of the 88,636 persons who tested negative, NAAT was performed in only 479 (0·54%). The median [IQR] antigen concentrations were 9·70 [4·98-34·11] pg/mL and 0·10 [0·01-0·19] pg/mL in the NAAT-positive and -negative samples, respectively. In specimens negative by NAAT, the frequency of high antigen concentrations monotonically approached zero, while NAAT positives did not follow any trend (Fig. 2).

**Figure 1.**
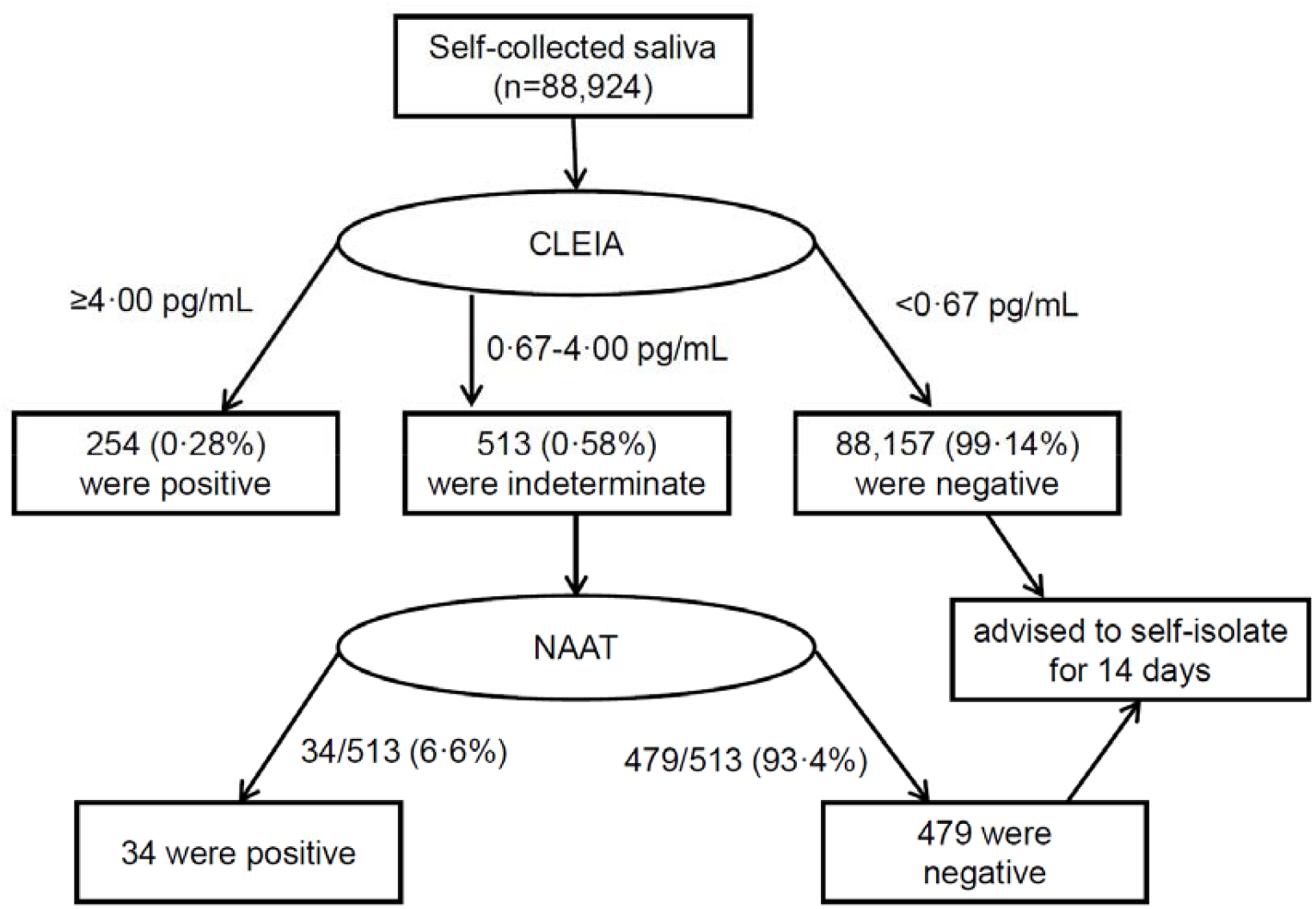
Flow chart of mass screening of international arrivals by the two-step strategy. 88,924 arrivals at international airports were screened using self-collected saliva. Initial CLEIA results were positive in 254 (0.28%) and negative in 88,157 (99.14%) persons. Confirmatory NAAT was only performed for samples in the indeterminate range (n=513; 0.58%).

**Figure 2.**
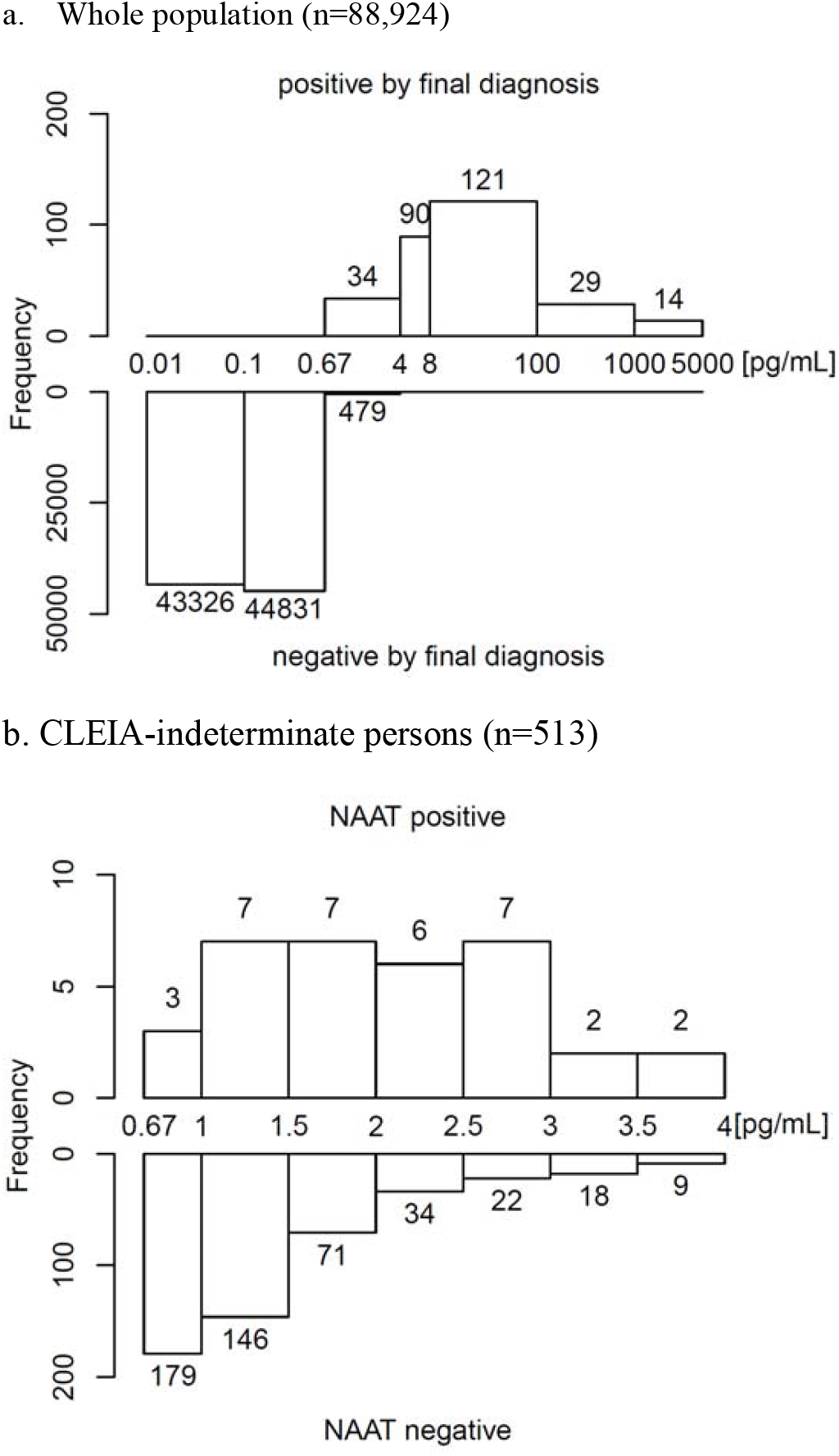
Barplots of viral antigen concentrations. (a) The frequency of viral antigen concentrations of the entire test population sorted by final diagnosis by the two-step strategy (288 positives and 88,636 negatives). (b) The frequency of antigen concentration in 513 persons judged to be indeterminate by initial CLEIA. NAAT was only performed for CLEIA results with antigen concentrations between 0.67 and 4.0pg/mL. The frequency of NAAT negative samples consistently approach zero with increasing antigen concentrations, while NAAT positives did not show any trend.

Comparing the effectiveness of the three strategies, the number of false negatives was greater in the two-step strategy compared to NAAT only in all scenarios of NAAT positivity, although both tests reduced false negative rates by more than half compared to test-free entry (Table 1). Conversely, the two-step strategy allowed the reduction of NAAT by approximately 95% compared to when NAAT was used in all persons. For example, in the scenario when *p*=0.1%, the NAAT only strategy detected 40 false negatives by 100,000 NAATs, whereas the two-step strategy resulted in 64 false negatives but only required 549 NAATs to be performed. Furthermore, as the majority of CLEIA indeterminates were NAAT negative, the number of NAATs needed did not significantly increase with varying scenarios of *p*.

**Table 1.** The effectiveness of three mass-screening strategies in a test population of 100,000 persons (when *FN/Pos*\*=0.4). The two-step strategy reduced the number of NAATs performed by approximately 95% compared to NAAT only, with an increase in false negatives by only 24 per 100,000 persons. Both NAAT only and two-step performed significantly better than free entry at limiting the number of false negatives.

| $p$ | NAAT only | | | Two-step<sup>†</sup> | | | Free entry | | |
| --- | --- | --- | --- | --- | --- | --- | --- | --- | --- |
|  | NAAT | Pos | FN | NAAT | Pos | FN | NAAT | Pos | FN |
| 0.1% | 100,000 | 100 | **40** | 549<br>[548-550] | 76<br>[68-83] | **64**<br>[57-72] | 0 | 0 | **140** |
| 0.2% | 100,000 | 200 | **80** | 558<br>[556-559] | 152<br>[136-166] | **128**<br>[114-144] | 0 | 0 | **280** |
| 0.5% | 100,000 | 500 | **200** | 583<br>[579-587] | 380<br>[340-415] | **320**<br>[285-360] | 0 | 0 | **700** |
| 1.0% | 100,000 | 1,000 | **400** | 626<br>[617-634] | 760<br>[680-830] | **640**<br>[570-720] | 0 | 0 | **1,400** |

$FN/Pos$ : ratio of false negatives to positives; $p$ : proportion of NAAT positivity; NAAT: number of nucleic acid amplification test performed; Pos: number of positive results; FN: number of false positives
\* $FN/Pos$ is the ratio of infected persons who test negative to all test positives.
<sup>†</sup> Estimated when the probability of CLEIA-positivity given NAAT-positivity is 76% in point estimates (90% credible interval between 68% to 83%).

When *FN/Pos* was set at 0.4, 0.1, and 2.0 in the scenario where *p* = 0.1%, the ratio of false negatives by NAAT only: two-step: free entry was 40 : 64 : 140 (i.e. 1 : 1.6 : 3.5), 100 : 124 : 200 (i.e. 1 : 1.2 : 2.0), and 200 : 224 : 300 (i.e. 1 : 1.1 : 1.5), respectively, showing increasingly diminished relative difference in efficacy between the three screening strategies (Table 2). Regardless of *FN/Pos*, post-screening 14-day quarantine substantially reduced imported false negatives, although the efficacy of quarantine was highly dependent on the degree of adherence (Table 2).

**Table 2.** Imported false negative (IFN) cases adjusted by 14-day quarantine in various settings of *FN/Pos*. Combining post-screening qurantine further decreased the number of imported cases false negatives depending on the adherence rate. The operational sensitivity of any strategy diminished with greater values of *FN/Pos*, with converging relative differences between the three strategies.

| Prob. of<br>NAAT<br>positivity | 14-days<br>Quarantine<br>Adherence | IFNs / 100,000 persons<br>(when $FN/Pos = 0.4$)* | | | IFNs / 100,000 persons<br>(when $FN/Pos = 1.0$)* | | | IFNs / 100,000 persons<br>(when $FN/Pos = 2.0$)* | | |
| --- | --- | --- | --- | --- | --- | --- | --- | --- | --- | --- |
|  |  | NAAT<br>only | Two-step<sup>†</sup> | Free<br>entry | NAAT<br>only | Two-step<sup>†</sup> | Free<br>entry | NAAT only | Two-step<sup>†</sup> | Free<br>entry |
| 0.1% | 0% | 40 | 64 [55-74] | 140 | 100 | 124 [117-132] | 200 | 200 | 224 [217-232] | 300 |
|  | 25% | 30 | 48 [41-56] | 105 | 75 | 93 [88-99] | 150 | 150 | 168 [163-174] | 225 |
|  | 50% | 20 | 32 [28-37] | 70 | 50 | 62 [59-66] | 100 | 100 | 112 [109-116] | 150 |
|  | 75% | 10 | 16 [14-19] | 35 | 25 | 31 [29-33] | 50 | 50 | 56 [54-58] | 75 |
| 0.2% | 0% | 80 | 128 [110-148] | 280 | 200 | 248 [234-264] | 400 | 400 | 448 [434-464] | 600 |
|  | 25% | 60 | 96 [83-111] | 210 | 150 | 186 [176-198] | 300 | 300 | 336 [326-348] | 450 |
|  | 50% | 40 | 64 [55-74] | 140 | 100 | 124 [117-132] | 200 | 200 | 224 [217-232] | 300 |
|  | 75% | 20 | 32 [28-37] | 70 | 50 | 62 [59-66] | 100 | 100 | 112 [109-116] | 150 |
| 0.5% | 0% | 200 | 320 [275-370] | 700 | 500 | 620 [585-660] | 1000 | 1000 | 1120 [1085-1160] | 1500 |
|  | 25% | 150 | 240 [206-278] | 525 | 375 | 465 [439-495] | 750 | 750 | 840 [814-870] | 1125 |
|  | 50% | 100 | 160 [138-185] | 350 | 250 | 310 [293-330] | 500 | 500 | 560 [543-580] | 750 |
|  | 75% | 50 | 80 [69-93] | 175 | 125 | 155 [146-165] | 250 | 250 | 280 [271-290] | 375 |
| 1.0% | 0% | 400 | 640 [550-740] | 1400 | 1000 | 1240 [1170-1320] | 2000 | 2000 | 2240 [2170-2320] | 3000 |
|  | 25% | 300 | 480 [413-555] | 1050 | 750 | 930 [878-990] | 1500 | 1500 | 1680 [1628-1740] | 2250 |
|  | 50% | 200 | 320 [275-370] | 700 | 500 | 620 [585-660] | 1000 | 1000 | 1120 [1085-1160] | 1500 |
|  | 75% | 100 | 160 [138-185] | 350 | 250 | 310 [293-330] | 500 | 500 | 560 [543-580] | 750 |
| any | 100% | 0 | 0 | 0 | 0 | 0 | 0 | 0 | 0 | 0 |

$FN/Pos$ : ratio of false negatives to positives; NAAT: nucleic acid amplification test
\* $FN/Pos$ is the ratio of infected persons who test negative to all test positives.
<sup>†</sup> Estimated when the probability of CLEIA-positivity given NAAT-positivity is 76% in point estimates (90% credible interval between 68% to 83%).

## Discussion

Although PCR is a standard of care for the detection of SARS-CoV-2, mandatory mass-screening should ideally avoid time-consuming and labor-intensive procedures. In this regard, quantitative antigen test by CLEIA is rapid albeit with slightly less accuracy than PCR^17^. Therefore, our two-step strategy combined the utility of initial CLEIA with the accuracy of NAAT only for diagnosis of indeterminate cases. This two-step strategy has been adopted in quarantine stations at the international airports in Japan, especially for the prevention of long waiting times spent in closed spaces in crowds and close-contact situations. Here, we showed the benefits of this strategy using 88,924 samples, providing numerical estimates of undetected infectees under various circumstances. The quantitative antigen testing allows for setting appropriate positive and negative thresholds to freely define the indeterminate range with a trade-off; a wider range improves test performance but at the expense of increasing the requirement of confirmatory NAAT. These thresholds may be altered to suit different clinical situations^19^, most importantly the local prevalence of disease.

Initial CLEIA has excellent specificity with the upper cutoff value at 4·00 pg/mL, as increasing antigen concentrations of NAAT negative samples consistently approach zero (Fig. 2b). Assuming that the frequency continues to decrease by one for every 0·5 pg/mL, 36 NAAT-negative samples would be included between 4 pg/mL and 8 pg/mL, giving specificity as high as 99·96% (=88,636/(88,636+36)). To further reduce the false positive rates, the upper cutoff may be set at higher values, but at the expense of increased requirement for confirmatory NAAT. For example, raising the upper cut-off from 4 pg/mL to 8 pg/mL would increase the number of indeterminate results and hence the number of NAAT from 513 to 603 (Fig. 2a). The main objective of screening for SARS-CoV-2 is to detect all transmissible persons, and this has been a great challenge with presymptomatic false negative PCR at 67% one day prior to and 38% on the day of symptom onset^20^. Therefore, in order to accurately identify presymptomatic infectees, pre-departure testing should be conducted several days before departure. Assuming that all passengers were asymptomatic with negative results before departure, infectees will most likely be in the latent phase at the time of screening. Given the median incubation period of 5 days^21-23^, testing five days prior to departure should reveal half the infections (i.e. *FN/Pos* ≃ 1.0) as false positives are very rare. Shorter intervals between pre-departure testing and inbound screening would yield more presymptomatic false negatives, and reduce operational test sensitivity^3, 20^. Illustrating this point, our results showed the ratio of false negatives comparing NAAT only to the two step strategy was 40 : 64 (i.e. 1 : 1.6) and 200 : 224 (i.e. 1 : 1.1) setting *FN/Pos* at 0.4 and 2.0, respectively. This trend was consistently seen in all scenarios of *p* and between any two strategies, indicating the vulnerability of depending on any test at a single time-point.

Regardless of testing strategy, post-screening quarantine performed very well at limiting import cases of false negatives in any scenario. However, perfect adherence to two weeks of compulsory isolation by all travelers is unrealistic, with detrimental psychological, social, and economic impact for those who do comply^24-26^. Recently, Wells et al. reported that testing on day 6 may reduce the duration of a 14-day quarantine by 50% while effectively preventing expected transmission events^27^. As with pre-departure, the timing of testing is essential as infected individuals very early in the incubation period may not be detected due to low viral loads. Nevertheless, mass screening at airport has benefits in reducing false negatives, especially in combination with well-timed pre-departure testing (i.e. when *FN/Pos* is small). Furthermore, screening is useful in monitoring the dynamics of test positivity, which may influence various immigration and health policies as well as suggest the possibility of viral mutations.

The main limitation of our study was the lack of clinical information after screening to assess the rates of observed false positivity. Post-screening investigation of more than 80,000 persons was simply out of the scope of this study. An additional limitation was that the probability of CLEIA-positive given NAAT-positive could not be validated, as NAAT was not performed for CLEIA-negative samples (antigen levels below 0·67pg/mL). Finally, although not truly a limitation of our study, we alluded to NAAT positivity as infectiousness, whereas this may not be true in cases of high Ct values^28, 29^.

In summary, we examined the data from mass screening of more than 88,000 persons at airport quarantines and showed the effectiveness of the two-step strategy. We believe the logistic advantage of reducing the burden of NAAT by approximately 95% far outweigh the cost of slightly higher imported cases of false negatives. Two-step testing by CLEIA followed by NAAT is effective in real-world situations, especially when combined with appropriately timed pre-departure testing and/or with quarantine optimized with repeat testing.

## Data Availability

Data may be available after consideration.

## Author Contributions

IY, PYS and TT provided statistical analysis and drafted the manuscript and all authors reviewed critically and approved the final manuscript.

## Reference

1 MA Honein, A Christie, DA Rose, et al. Summary of guidance for public health strategies to address high levels of community transmission of sars-cov-2 and related deaths, december 2020. MMWR Morb Mortal Wkly Rep 2020; 69(49):1860–1867.

2 JT Wu, K Leung, GM Leung. Nowcasting and forecasting the potential domestic and international spread of the 2019-ncov outbreak originating in wuhan, china: A modelling study. Lancet 2020; 395(10225):689–697.

3 M Chinazzi, JT Davis, M Ajelli, et al. The effect of travel restrictions on the spread of the 2019 novel coronavirus (covid-19) outbreak. Science 2020; 368(6489):395–400.

4 JY Liu, TJ Chen, SJ Hwang. Analysis of imported cases of covid-19 in taiwan: A nationwide study. Int J Environ Res Public Health 2020; 17(9).

5 BL Dickens, JR Koo, JT Lim, et al. Strategies at points of entry to reduce importation risk of covid-19 cases and re-open travel. J Travel Med 2020.

6 M Bielecki, D Patel, J Hinkelbein, et al. Air travel and covid-19 prevention in the pandemic and peri-pandemic period: A narrative review. Travel Med Infect Dis 2020; 39(101915.

7 LH Chen, R Steffen. Sars-cov-2 testing to assure safety in air travel. J Travel Med 2021.

8 N Sethuraman, SS Jeremiah, A Ryo. Interpreting diagnostic tests for sars-cov-2. JAMA 2020; 323(22):2249–2251.

9 W Wang, Y Xu, R Gao, et al. Detection of sars-cov-2 in different types of clinical specimens. JAMA 2020; 323(18):1843–1844.

10 ML Bastos, S Perlman-Arrow, D Menzies, JR Campbell. The sensitivity and costs of testing for sars-cov-2 infection with saliva versus nasopharyngeal swabs: A systematic review and meta-analysis. Ann Intern Med 2021.

11 TS Higgins, AW Wu, J. Ting. Sars-cov-2 nasopharyngeal swab testing-false-negative results from a pervasive anatomical misconception. JAMA Otolaryngol Head Neck Surg 2020.

12 AL Wyllie, J Fournier, A Casanovas-Massana, et al. Saliva or nasopharyngeal swab specimens for detection of sars-cov-2. N Engl J Med 2020; 383(13):1283–1286.

13 L Caulley, M Corsten, L Eapen, et al. Salivary detection of covid-19. Ann Intern Med 2020.

14 I Yokota, PY Shane, K Okada, et al. Mass screening of asymptomatic persons for sars-cov-2 using saliva. Clin Infect Dis 2020.

15 ME Kretzschmar, G Rozhnova, MCJ Bootsma, et al. Impact of delays on effectiveness of contact tracing strategies for covid-19: A modelling study. Lancet Public Health 2020; 5(8):e452–e459.

16 Y Hirotsu, M Maejima, M Shibusawa, et al. Comparison of automated sars-cov-2 antigen test for covid-19 infection with quantitative rt-pcr using 313 nasopharyngeal swabs, including from seven serially followed patients. Int J Infect Dis 2020; 99(397–402.

17 I Yokota, PY Shane, K Okada, et al. A novel strategy for sars-cov-2 mass-screening using quantitative antigen testing of saliva. SSRN Electronic Journal 2020.

18 M Al-Qahtani, S AlAli, A AbdulRahman, et al. The prevalence of asymptomatic and symptomatic covid-19 in a cohort of quarantined subjects. Int J Infect Dis 2020; 102(285-288.

19 S Woloshin, N Patel, AS Kesselheim. False negative tests for sars-cov-2 infection - challenges and implications. N Engl J Med 2020; 383(6):e38.

20 LM Kucirka, SA Lauer, O Laeyendecker, et al. Variation in false-negative rate of reverse transcriptase polymerase chain reaction-based sars-cov-2 tests by time since exposure. Ann Intern Med 2020; 173(4):262–267.

21 SA Lauer, KH Grantz, Q Bi, et al. The incubation period of coronavirus disease 2019 (covid-19) from publicly reported confirmed cases: Estimation and application. Annals of Internal Medicine 2020; 172(9):577–582.

22 H-Y Cheng, S-W Jian, D-P Liu, et al. Contact tracing assessment of covid-19 transmission dynamics in taiwan and risk at different exposure periods before and after symptom onset. JAMA Internal Medicine 2020; 180(9):1156–1163.

23 NM Linton, T Kobayashi, Y Yang, et al. Incubation period and other epidemiological characteristics of 2019 novel coronavirus infections with right truncation: A statistical analysis of publicly available case data. J Clin Med 2020; 9(2).

24 SK Brooks, RK Webster, L. Smith, et al. The psychological impact of quarantine and how to reduce it: Rapid review of the evidence. Lancet 2020; 395(10227):912–920.

25 S Dubey, P Biswas, R Ghosh, et al. Psychosocial impact of covid-19. Diabetes Metab Syndr 2020; 14(5):779–788.

26 VJ Clemente-Suarez, AA Dalamitros, AI Beltran-Velasco, et al. Social and psychophysiological consequences of the covid-19 pandemic: An extensive literature review. Front Psychol 2020; 11(580225.

27 CR Wells, JP Townsend, A Pandey, et al. Optimal covid-19 quarantine and testing strategies. Nat Commun 2021; 12(1):356.

28 J Bullard, K Dust, D Funk, et al. Predicting infectious sars-cov-2 from diagnostic samples. Clin Infect Dis 2020.

29 B La Scola, M Le Bideau, J Andreani, et al. Viral rna load as determined by cell culture as a management tool for discharge of sars-cov-2 patients from infectious disease wards. Eur J Clin Microbiol Infect Dis 2020; 39(6):1059–1061.

